# The association between maternal parenting styles and eating disorders among Lebanese university students: The mediating role of the self-assessment dimension of self-esteem

**DOI:** 10.1101/2025.02.06.25321782

**Authors:** Toni Sawma, Myriam El Khoury-Malhame, Tatiana Hayek, Jana Mourad, Sandrella Bou Malhab, Rana Rizk

**Affiliations:** Department of Psychology and Education, School of Arts and Sciences, Lebanese American University, Byblos, Lebanon; Department of Nutrition and Food Science, School of Arts and Sciences, Lebanese American University, Byblos, Lebanon; Institut National de Santé Publique, d’Épidémiologie Clinique et de Toxicologie-Liban (INSPECT-LB), Beirut, Lebanon

**Keywords:** Eating disorders, Eating behaviors, Maternal parenting styles, Self-esteem, Self-assessment, University students, Lebanon

## Abstract

**Objective:** Eating disorders are influenced by an array of distinct factors such as genetics, personality traits, and history of childhood abuse. Recently maternal parenting styles have been investigated as one of the contributing aspects. These styles are intricately connected to children’s self-esteem. As such this study aims to examine the association between maternal parenting styles and eating disorders and better investigate the mediating role of the self-assessment, an essential understudied dimension of self-esteem.

**Method:** A cross-sectional online study was conducted between November 2023 and May 2024. Around 300 Lebanese university students filled out surveys assessing demographic factors, maternal parenting style, eating disorders, and self-assessment.

**Results:** We found that different maternal styles distinctly influence the development of eating disorders; with self-esteem mediating the relationship between different maternal parenting styles and eating disorders development. Self-assessment negatively mediated the association between maternal expectations and eating control and dieting, and it negatively mediated the association between maternal permissive and dieting and bulimia. Self-assessment positively mediated the association between maternal control and eating control, dieting, and bulimia.

**Discussion:** This study mostly provides healthcare professionals and clinical educators with valuable insights into the complex association between parenting and eating disorders and highlights the importance of developing skillsets to boost self-esteem for those at risk of developing eating disorders.

## Introduction

Eating disorders (ED), such as anorexia nervosa and bulimia nervosa, are a broad spectrum of complex mental health conditions characterized by unhealthy eating patterns and distorted body image (Culbert et al.,2015). ED can be accompanied by extreme disabling preoccupation and distress concerning one’s body weight leading to an impairment in overall well-being and adverse physical and psychological consequences including malnutrition, heart problems, depression, and anxiety (Culbert et al.,2015; Keski-Rahkonen & Mustelin, 2016). Globally, the prevalence of ED oscillates between 2.58% - 8.4% for females and 0.74%-2.2% for males and seems to follow a worrisome exponential trend in many societies (Hay et al., 2023). ED has been reportedly caused by different clusters of factors including biological, psychological and social ones. Traditionally, genetics, neurotransmitters, or hormonal imbalances have been on the forefront of the link to the progression of ED (Herpertz Dahlmann, 2015). More recently, additional contributing factors have been investigated, such as perfectionism, family dynamics, childhood trauma as well as a cultural obsession with thinness and social pressure to conform (Hilbert et al., 2014; Stice et al., 2020).

## Theoretical Framework

The sociocultural model developed by Neumark-Sztainer et al. (1996) is based on the social cognitive theory (SCT) postulating that socio-environmental and cultural factors’ influence on behavioral aspects are facilitated by their impact on personal factors. In our study, the model posits that the interaction among different personal, socio-environmental cultural, and behavioral factors result in maladaptive eating behaviors that progress into ED. This is mostly because the socio-environmental and cultural factors contribute to the internalization of thin ideals with social comparison and body surveillance contributing to body dissatisfaction and the development of an ED such as anorexia nervosa and bulimia nervosa (Saunders & Eaton, 2018). Furthermore, cognitions act as mediators between the individuals’ behaviors and the environment in accordance to family dynamics. In that framework, family factors have been increasingly studied about ED. Dysfunctional family dynamics could contribute to increased ED (carnelian et al.2017), whereas adequate parenting styles, involving synchrony of control and warmth, is a major protective factor (Hampshire et al.,2022; Ningning & Wenguang, 2023; Ramsewak et al.,2022). Thus, parenting styles additionally influence how individuals internalize sociocultural messages and express them in their daily lives, and in this regard, individual self-esteem could mediate this relationship (Kroplewski et al., 2019; Peng et al.,2021; Theodoropoulou et al., 2023).

Parenting styles are generally clustered according to 2 variables: levels of control and warmth, into authoritarian, neglectful, avoidant, and authoritative styles accordingly (Pinquart et al.,2017). Accumulating evidence shows that all except authoritative positively correlate with the development of ED (Khosravi et al.,2023). As such, authoritarian, permissive, and neglectful parenting style could increase children’s proneness to develop an ED, especially during adolescence (Usmani et al., 2022). In this context, data also points to a significant differential contribution from maternal and paternal styles to ED (Sahota et al. 2024). This is particularly relevant in collectivist Levantine cultures as mothers are tasked more frequently with the care of children. According to the social learning theory, mothers can reinforce unhealthy relations to food as their children might be learning and acquiring food-related behaviors by observing and imitating them (Bandura, 1986, as cited in Fryling et al., 2011).

The impact of mothers on their children’s development of ED is far from homogenous to all siblings and seems additionally mediated by individual factors such as gender and self-esteem (Peng et al.,2021; Theodoropoulou et al.,2023). Interestingly, subdivisions of self-esteem have been shown to differentially influence and lower the development of ED (Bardone-Cone et al.,2020; Paranjothy & Wade, 2024). Self-esteem is indeed a multifaceted construct and tackling its subsets might provide more scientific and clinical usefulness as for instance it can help identify which subset specifically mediates the relation between parenting attitudes and disordered eating and explain how specific practices can protect from maladaptive patterns. These subdivisions include first self-assessment or the positive evaluation of personal qualities, and second self-acceptance or not judging oneself. Self-assessment is one such essential dimension as it reflects an individual’s appraisal of their own abilities, worth, and competence. It plays a role in shaping thoughts, feelings and behaviors. Self-assessment has been found to act as a direct mediator in the relation between parenting styles and ED, especially the maternal one (Kroplewski et al., 2019; Peng et al.,2021). It could be that authoritative parenting, characterized by warmth, responsiveness, and clear expectations, can promote positive self-assessment and in turn make children more resilient to negative body image and subsequently decrease risks of eating disorders.

Given the worrisome rising trend of ED notably among university students in the Middle East (Melisse et al., 2020), and the increased interest in maternal parenting style, our study aims at investigating the protective effect of authoritative mother parenting on eating disorders working through its influence on self-assessment. In fact, the rise in food insecurity, social media pressure, the “thin ideal” belief, and cultural encouragement of dieting practices globally (Rana et al., 2023; Kharroubi et al., 2021; Melisse et al., 2020), and the particularly close mother-daughter relationship locally, accounting for ED could be buffered by maternal parenting style whereby this protective factor could dampen young adults dissatisfaction with appearance and subsequent eating concerns, (Azzeh et al., 2022; Deek et al., 2024; Tavolacci et al., 2021). This relation remains particularly understudied among university students, notably those living in adverse situations with scarce resources such as those in Lebanon. Lebanon is a Levantine country with a patriarchal collectivist society, and has been navigating massive protracted crises since the COVID-19 pandemic, including sociopolitical unrest and the devaluation of local currency (El Khoury-Malhame et al., 2023). This has severely exacerbated rates of overall mental distress including ED, especially among university students (Abouzeid et al., 2021; Deek et al., 2024). To better address this lack of research in that regard, examining the association between different maternal parenting styles and various subsets of ED becomes crucial for addressing this rising public health issue. We also explore the impact of self-assessment, a specific dimension of self-esteem, on this association. Additionally, the body mass index (BMI) was examined due to previous research showing that it has a predictive role and an important added value when exploring mediators and parenting styles’ influence on eating disorders (Abdulkadir et al., 2020; Pruccoli et al.,2023). In line with the theoretical framework, we hypothesized that self-assessment will mediate the relation between authoritative maternal parenting style and lower levels of ED.

## Methodology

### Ethical considerations

Ethical approval was obtained from the Lebanese American University Institutional Review Board (LAU IRB: LAU.SAS.TS1.10/Nov/2023). Data collection was conducted anonymously. Before completing the questionnaire, all participants provided their written consent by clicking on the respective button on the first page of the Google Form.

### Study design

This cross-sectional study utilized an online survey administered via Google Forms. The questionnaire, developed in English, was distributed through social media platforms (WhatsApp, Facebook and Instagram) and via a QR code. Participants were also encouraged to share the survey link with their classmates and friends. Data was collected between November 15 2023 and December 15, 2023.

### Study population

The total study population consisted of around 300 university students, aged 18-25 years. Only participants who provided consent were included.

### Sample size calculation

The sample size was determined using G*Power, version 3.1 based on an effect size of 0.22, an alpha level of 0.05, and a power of 0.95, which indicated a required sample of 258 students. The effect size was derived from a correlation coefficient of 0.11 between an authoritarian maternal parenting style and ED symptomatology, as reported by Kaufman (2015). We deliberately selected the smallest correlation coefficient from this study to maximize the required sample size. The effect size of 0.22 was calculated using the Pearson correlation coefficient of 0.11 following the method outlined by Becker (2000). To account for potential non-completion of the questionnaire, the calculated sample size was increased by 10%, resulting in a final target of 283 participants.

### Data collection tool

The first section of the questionnaire included socio-demographics (age, gender, governorate, living arrangements, number of people living in the dwelling, and number of rooms), as well as self-reported weight and height. The crowding index was calculated by dividing the number of residents by the number of rooms and served as a proxy for socioeconomic status. BMI was calculated by dividing reported weight in kilograms by reported height in meters squared.

#### Mother’s Parental Authority Questionnaire (PAQ)

Buri (1989, 1991). It assesses parental authority prototypes including permissive parenting characterized by low control and maturity demands but high communication and responsiveness, authoritarian parenting characterized by high control and maturity demands but low responsiveness and communication, and authoritative parenting characterized by high levels of control, responsiveness, communication, and maturity demands. The PAQ consists of 30 items designed to assess participants’ retrospective perceptions of their mother’s parenting styles during their formative years. Respondents indicated their level of agreement with statements related to various maternal parenting behaviors, including expectations, decision-making, disciplinary methods, and communication styles, using a 5-point Likert scale ranging from Strongly Disagree to Strongly Agree. Higher scores reflect greater alignment with a given parental prototype.

**Eating Attitudes Test (EAT)-26**, a screening tool designed to help individuals assess the potential presence of an ED that may require professional attention. The EAT-26 does not provide a definitive diagnosis but serves as a preliminary measure to guide individuals toward seeking professional consultation. The questionnaire comprises 26 items, each with six response options ranging from Never to Always. It yields three factors: the Dieting Subscale which relates to avoidance of fattening foods and preoccupation with being thinner; Bulimia, which concerns thoughts about food and symptoms indicative of bulimia; and the eating control, which involves self-control of eating and perceived pressure from others to gain weight. For each factor, a score is calculated by summing the responses, with higher scores suggesting more pronounced disordered eating attitudes.

#### Rosenberg Self-Esteem Scale, Self-Assessment (SA) Dimension

It comprises five items from the original scale and invites assessment of qualities, reflecting self-competence. The instrument demonstrates good psychometric properties (Rosenberg, 1965; Tafarodi et al.,2002). Respondents indicated their level of agreement or disagreement with each statement using a 4-point Likert scale ranging from Strongly Disagree to Strongly Agree.

#### Body Maxx index (BMI)

The BMI was calculated based on reported weight and height in addition to measuring it with the Quetelet index. To elaborate, the weight was divided by the height squared (W/H 2 [kg/m 2]).

### Statistical analysis

Data from completed forms was imported into a Microsoft Excel spreadsheet. Data analysis was then performed on SPSS software version 25 (Chicago, IL, USA). A descriptive analysis was performed using absolute frequencies and percentages for categorical variables and means and standard deviations (SD) for quantitative measures. Pearson’s correlation was used for bivariate analysis. Assumptions of continuous variables normality and other conditions were checked via histogram and normal probability plot.

Construct validity of the scales was assessed using the rotated component matrix technique. The Kaiser-Meyer-Olkin measure of sampling adequacy and Bartlett’s test of sphericity were calculated to ensure the model’s adequacy. Factors with eigenvalues >1 were retained, and the scree plot method, which is the Ellis JL. Factor and item analysis was used to determine the number of components to extract. Only items with factor loading > 0.4 were considered. Moreover, the internal consistency of the scales was assessed using Cronbach’s alpha.

A mediation analysis using PROCESS version 4.2 was used to calculate three pathways in the mediation analysis. While pathway A determined the regression coefficient for the effect of maternal expectations and control scales on self-assessment scale, pathway B examined the association between the self-assessment on bulimia, control, and dieting scales and pathway C estimated the total and direct effect of maternal expectations and control scales on bulimia, control, and dieting scales. The Macro generated bias-corrected bootstrapped 95% confidence intervals (CI) to test the significance of the indirect effect. Mediation was considered significant when the CI for the indirect effect did not include zero. A p-value less than 0.05 was considered statistically significant. (Figure 1) The covariates that were included in the mediation model were those that showed associations with p value <0.2 in the bivariate analysis. A p-value less than 0.05 was considered significant.

**Figure 1.**
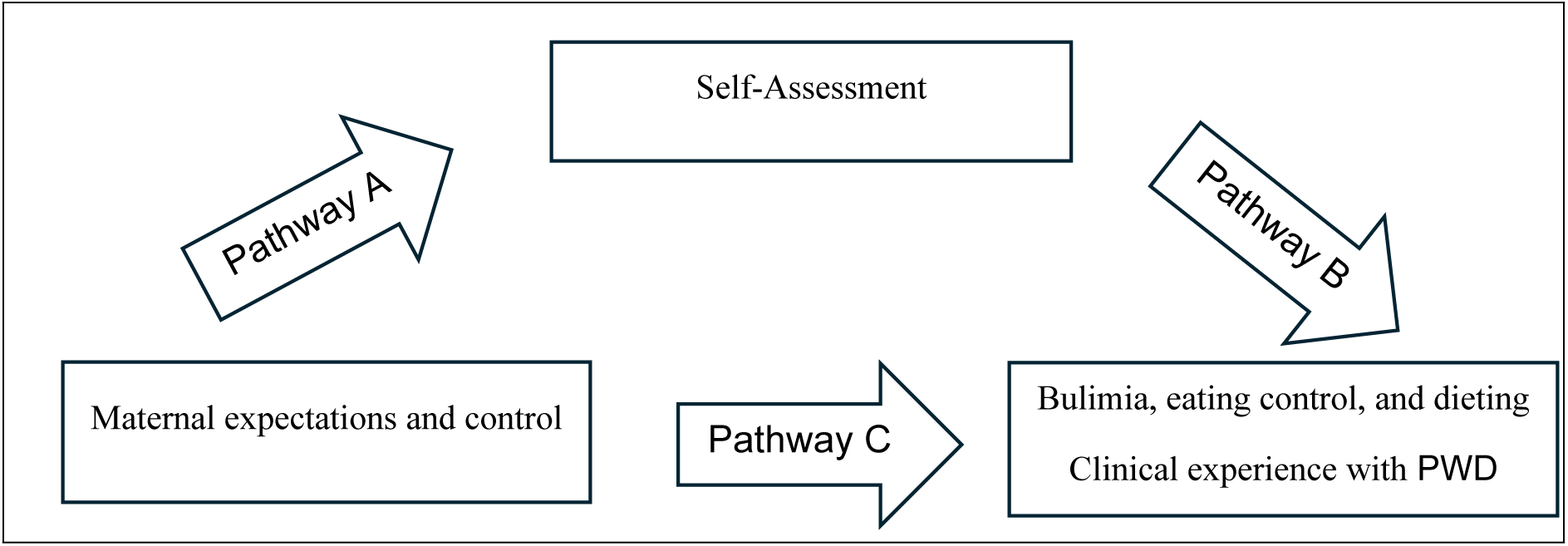
Mediation framework – assessment of the mediation of the self-assessment on the association between maternal expectations and control and bulimia, eating control, and dieting

**Figure 1.**
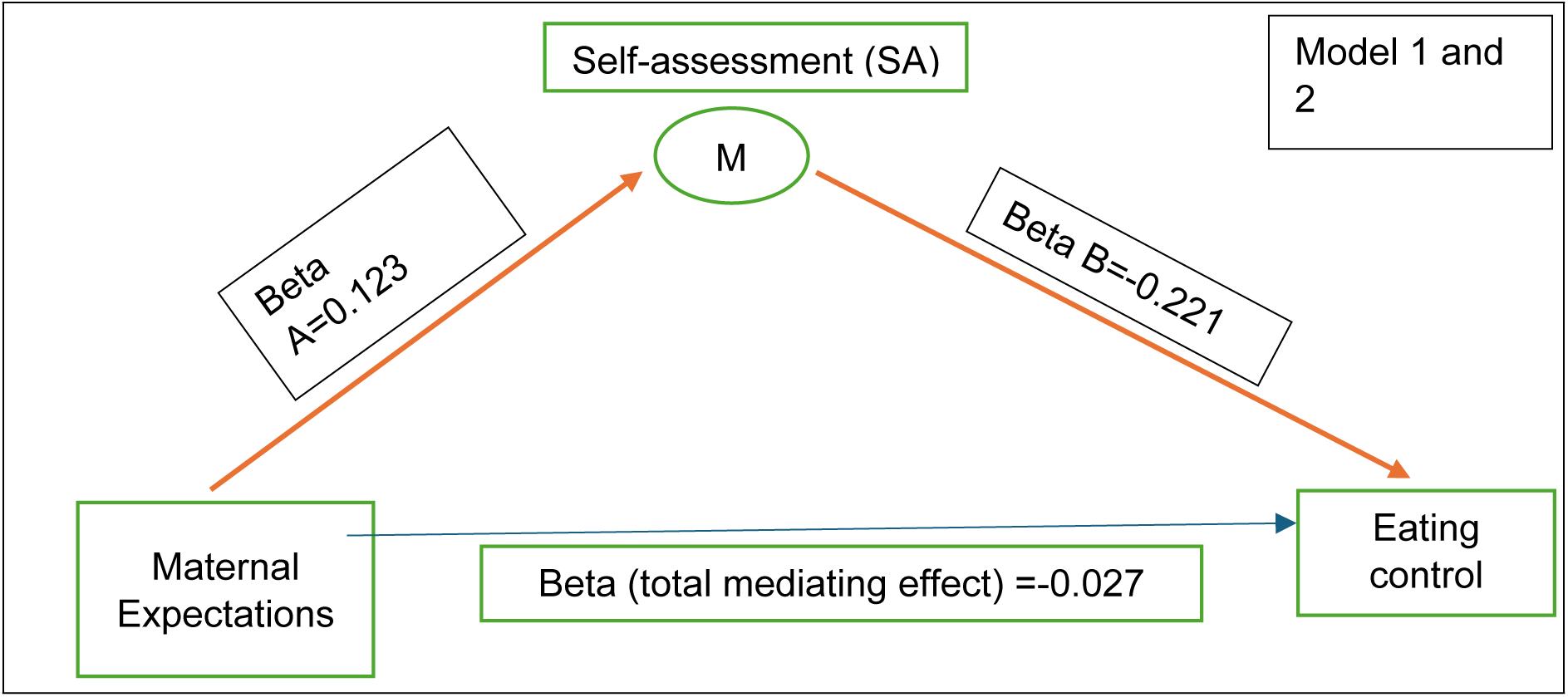
Mediating effect of Self-Assessment (SA) dimension on eating control in terms of maternal expectations (Red bold arrows illustrate the significant relationships)

## Results

### Sample characteristics

In total, 299 participants were included. Most of the sample were females (68.9%), with a mean age of 21 years and a mean BMI of 23.15 Kg/m^2^.

### Factor analysis. Construct validity and reliability of the “maternal” scales

Firstly, all the items used by the authors of the three used tools were included in the factor analysis of the total scales. This process led to factor structures that lacked coherence or comprehensibility and could not explain the objective of the scales’ construction, in addition to inadequate loading on respective factors. Therefore, for each of the scales, each subscale was considered as an independent entity and underwent a separate factor analysis.

For each scale, the subscales validated by the factor analysis converged adequately and confirmed the contents of the initial tools constructed by the authors of the initial tools although yielding a different factor structures and composition (Appendix1).

### Bivariate analysis

Table 2. showed that self-assessment (SA) was positively and significantly correlated with maternal expectations and permissive, while negatively correlated with eating control. Eating Control Subscale was significantly and negatively correlated with self-assessment and maternal expectations, and positively correlated with maternal control. Dieting was significantly and negatively correlated with self-assessment and maternal permissive while positively correlated with maternal control and expectations. Finally, bulimia was significantly and negatively correlated with self-assessment and maternal expectations while positively correlated with maternal control.

**Table 1.** Sample characteristics (n=299)

| Table 1. Sample characteristics (n=299) |  |
| --- | --- |
| Variable | N (%) |
| Gender |  |
| Male | 90 (31.1%) |
| Female | 199 (68.9%) |
|  | **Mean (SD)** |
| Age (years) | 21.08 (2.83) |
| BMI | 23.15 (4.28) |
| Self-Assessment (SA) dimension | 12.43 (2.94) |
| Maternal expectations | 30.35 (7.38) |
| Maternal permissive | 22.78 (4.97) |
| Maternal control | 19.73 (6.12) |
| Bulimia | 2.73 (3.67) |
| Eating Control | 2.65 (3.44) |
| Dieting | 9.36 (8.72) |

**Table 2.** Bivariate analysis.

| **Table 2. Bivariate analysis** |  |  |  |  |  |  |  |  |
| --- | --- | --- | --- | --- | --- | --- | --- | --- |
|  | Self-Assessment |  | Eating Control |  | Dieting |  | Bulimia |  |
|  | r | P value | r | P value | r | P value | r | P value |
| Self-Assessment |  |  | -0.213 | 0.001* | -0.230 | <0.001* | -0.183 | 0.004* |
| Maternal expectations | 0.320 | <0.001* | -0.158 | 0.008* | 0.196 | 0.001* | -0.228 | <0.001* |
| Maternal control | -0.183 | 0.003* | 0.130 | 0.03* | 0.191 | 0.001* | 0.200 | 0.001* |
| Maternal permissive | 0.120 | 0.055* | -0.038 | 0.531 | -0.164 | 0.006* | -0.064 | 0.262 |
| *Significant results |  |  |  |  |  |  |  |  |

Abbreviations: Self-Assessment (SA), Maternal expectations and control, bulimia, eating control, and dieting.

### Mediation analysis Mediation analysis summary

Table 3 presents the correlation between parenting styles and eating disorders, with mediation analysis involving the self-esteem. For each of the studied parenting styles, while the indirect effect on type of eating disorder was significant, the absence of a significant direct effect suggests full mediation.

**Table 3.** Mediation analysis.

| Table 3. Mediation analysis |  |  |  |  |  |  |
| --- | --- | --- | --- | --- | --- | --- |
| Mediating effect of Self-Assessment on eating Control in terms of maternal expectations |  |  |  |  |  |  |
| Model 1: Linear regression taking Self-Assessment as dependent variable and maternal expectations as the independent variable |  |  |  |  | Indirect effect of maternal expectations on eating control (SA as mediator) |  |
| Factor | US Beta | SD Beta | 95% CI) | P value |  |  |
| PAQM expectations | 0.123 | 0.025 | 0.073; 0.173 | <0.001 |  |  |
| Model 2: Linear regression taking eating Control as dependent variable and maternal expectations and Self-Assessment as independent variables |  |  |  |  | US Beta | 95% CI |
| Factor | US Beta | SD Beta | 95% CI) | P value | -0.027 | -0.049; -0.010 |
| PAQM expectations | -0.021 | -0.680 | -0.800; 0.039 | 0.497 |  |  |
| SA | -0.221 | -3.025 | -0.365; -0.077 | 0.003 |  |  |
| Mediating effect of Self-Assessment on eating Control in terms of maternal control |  |  |  |  |  |  |
| Model 3: Linear regression taking Self-Assessment as dependent variable and maternal control as the independent variable |  |  |  |  | Indirect effect of maternal control on eating control (SA as mediator) |  |
| Factor | US Beta | SD Beta | 95% CI) | P value |  |  |
| PAQM control | -0.089 | -2.860 | -0.151; -0.028 | 0.004 |  |  |
| Model 4: Linear regression taking eating control as dependent variable and maternal control and Self-Assessment as independent variables |  |  |  |  | US Beta | 95% CI |
| Factor | US Beta | SD Beta | 95% CI) | P value | 0.020 | 0.005; 0.044 |
| Maternal control | 0.001 | 0.020 | -0.068; 0.070 | 0.984 |  |  |
| SA | -0.036 | -3.320 | -0.376; -0.096 | 0.001 |  |  |
| Mediating effect of Self-Assessment (SA) on Dieting Subscale in terms of maternal expectations |  |  |  |  |  |  |
| Model 5: Linear regression taking Self-Assessment as dependent variable and maternal expectations as the independent variable |  |  |  |  | Indirect effect of maternal expectations |  |

| Factor | US Beta | SD Beta | 95% CI | P value | on Dieting Subscale (SA as mediator) |  |
| --- | --- | --- | --- | --- | --- | --- |
| Maternal expectations | 0.128 | 5.023 | 0.078; 0.179 | <0.001 |  |  |
| **Model 6: Linear regression taking Dieting Subscale as dependent variable and maternal expectations and Self-Assessment as independent variables** |  |  |  |  | US Beta | 95% CI |
| Factor | US Beta | SD Beta | 95% CI) | P value | -0.069 | -0.124; -0.027 |
| PAQM expectations | -0.102 | -1.361 | -0.246; 0.042 | 0.166 |  |  |
| SA | -0.535 | -3.045 | -0.880; -0.189 | 0.003 |  |  |
| **Mediating effect of Self-Assessment on Dieting Subscale in terms of maternal control** |  |  |  |  |  |  |
| **Model 7: Linear regression taking Self-Assessment as dependent variable and maternal control as independent variable** |  |  |  |  | **Indirect effect of maternal control on Dieting Subscale (SA as mediator)** |  |
| Factor | US Beta | SD Beta | 95% CI | P value |  |  |
| PAQM control | -0.094 | -2.980 | -0.157; -0.032 | 0.003 |  |  |
| **Model 8: Linear regression taking Dieting Subscale as dependent variable and maternal control and Self-Assessment as independent variables** |  |  |  |  | US Beta | 95% CI |
| Factor | US Beta | SD Beta | 95% CI) | P value | 0.054 | 0.015; 0.109 |
| PAQM control | 0.104 | 1.219 | -0.064; 0.271 | 0.224 |  |  |
| SA | -0.571 | -3.352 | -0.906; -0.235 | 0.001 |  |  |
| **Mediating effect of Self-Assessment on Dieting Subscale in terms of maternal permissive** |  |  |  |  |  |  |
| **Model 9: Linear regression taking Self-Assessment as dependent variable and maternal permissive as independent variable** |  |  |  |  | **Indirect effect of maternal permissive on Dieting Subscale (SA as mediator)** |  |
| Factor | US Beta | SD Beta | 95% CI | P value |  |  |
| PAQM permissive | 0.083 | 1.963 | -0.003; 0.166 | 0.05 |  |  |
| **Model 10: Linear regression taking Dieting as dependent variable and maternal permissive and Self-Assessment dimension as independent variables** |  |  |  |  | US Beta | 95% CI |
| Factor | US Beta | SD Beta | 95% CI) | P value | -0.049 | -0.115; -0.005 |
| PAQM permissive | -0.114 | -1.026 | -0.334; 0.105 | 0.306 |  |  |
| SA | -0.588 | -3.487 | -0.920; -0.256 | 0.006 |  |  |
| **Mediating effect of Self-Assessment on Bulimia in terms of maternal expectations** |  |  |  |  |  |  |
| **Model 11: Linear regression taking Self-Assessment as dependent variable and maternal expectations as independent variable** |  |  |  |  | **Indirect effect of maternal expectations on Bulimia (SA as mediator)** |  |
| Factor | US Beta | SD Beta | 95% CI | P value |  |  |
| PAQM expectations | 0.128 | 5.023 | 0.078; 0.179 | <0.001 |  |  |
| **Model 12: Linear regression taking Bulimia as dependent variable and maternal expectations and Self-Assessment dimension as independent variables** |  |  |  |  | US Beta | 95% CI |
| Factor | US Beta | SD Beta | 95% CI) | P value | -0.023 | -0.044; -0.006 |
| PAQM expectations | -0.046 | -1.460 | -0.108; 0.016 | 0.146 |  |  |
| SA | -0.178 | -2.359 | -0.326; -0.029 | 0.019 |  |  |
| **Mediating effect of Self-Assessment on Bulimia in terms of maternal control** |  |  |  |  |  |  |
| **Model 13: Linear regression taking Self-Assessment as dependent variable and maternal control as independent variable** |  |  |  |  | **Indirect effect of maternal control on Bulimia Subscale (SA as mediator)** |  |
| **Factor** | **US Beta** | **SD Beta** | **95% CI** | **P value** |  |  |
| PAQM control | -0.094 | -2.980 | -0.156; -0.032 | 0.003 |  |  |
| **Model 14: Linear regression taking Bulimia as dependent variable and maternal control and Self-Assessment as independent variables** |  |  |  |  | **US Beta** | **95% CI** |
| **Factor** | **US Beta** | **SD Beta** | **95% CI)** | **P value** | 0.019 | 0.004; 0.040 |
| Maternal control | 0.027 | 0.729 | -0.045; 0.099 | 0.467 |  |  |
| SA | -0.201 | -2.751 | -0.346; -0.057 | 0.006 |  |  |

### Analysis Summary

#### Mediating effect of Self-Assessment on Eating Control Subscale

Models 1 and 2 showed that self-assessment significantly and negatively mediated the association between maternal expectations and the eating control (total mediation). There was no significant direct effect of maternal expectations on eating control (Table 3; Figure 1).

Models 3 and 4 showed that self-assessment significantly and positively mediated the association between maternal control and eating control (Total mediation). There was no significant direct effect of maternal control on the eating control (Table 3; Figure 2).

**Figure 2.**
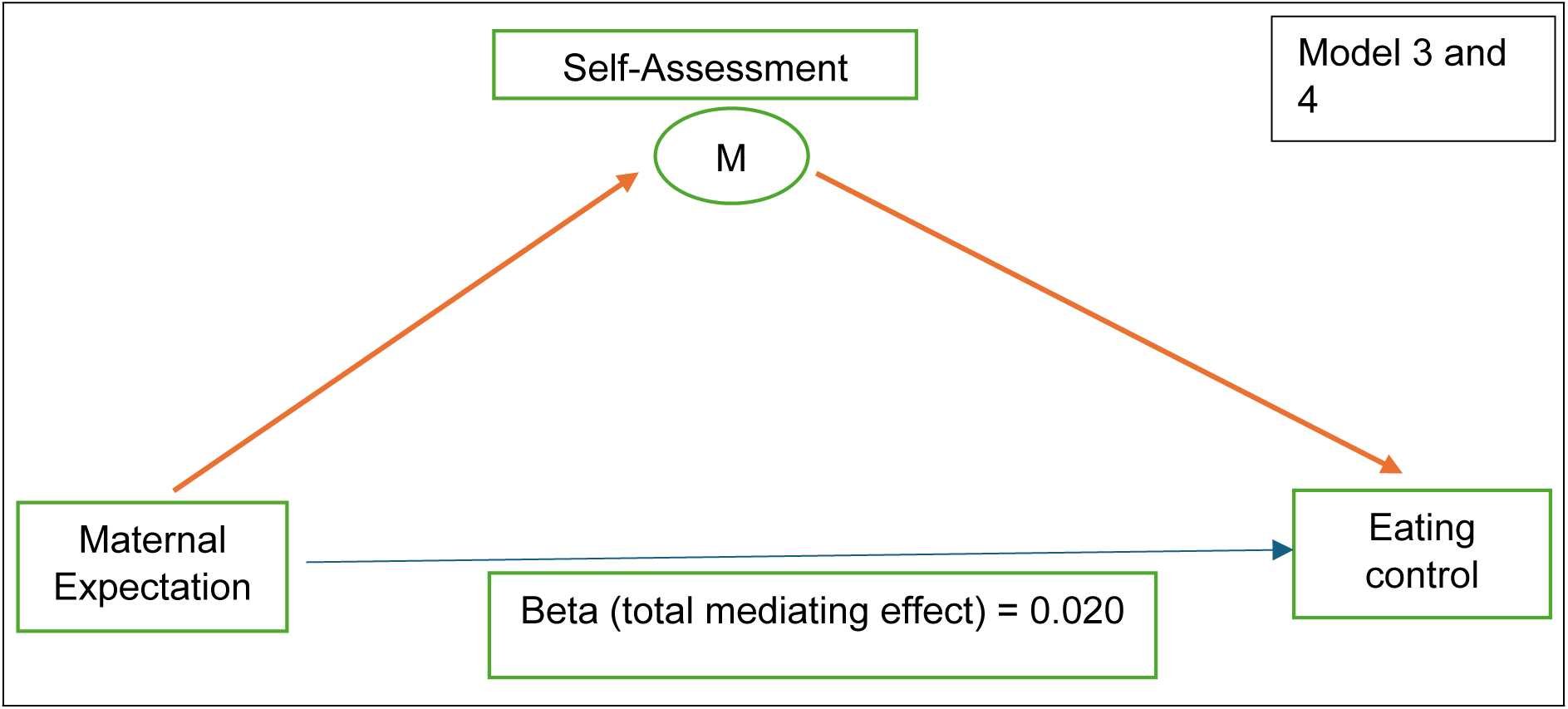
Mediating effect of Self-Assessment (SA) dimension on eating control in terms of maternal expectations (Red bold arrows illustrate the significant relationships)

#### Mediating Effect of Self-Assessment on Dieting Subscale

Models 5 and 6 showed that self-assessment significantly and negatively mediated the association between maternal expectations and dieting (Total mediation). There was no significant direct effect of maternal expectations on the dieting (Table 3; Figure 3).

**Figure 3.**
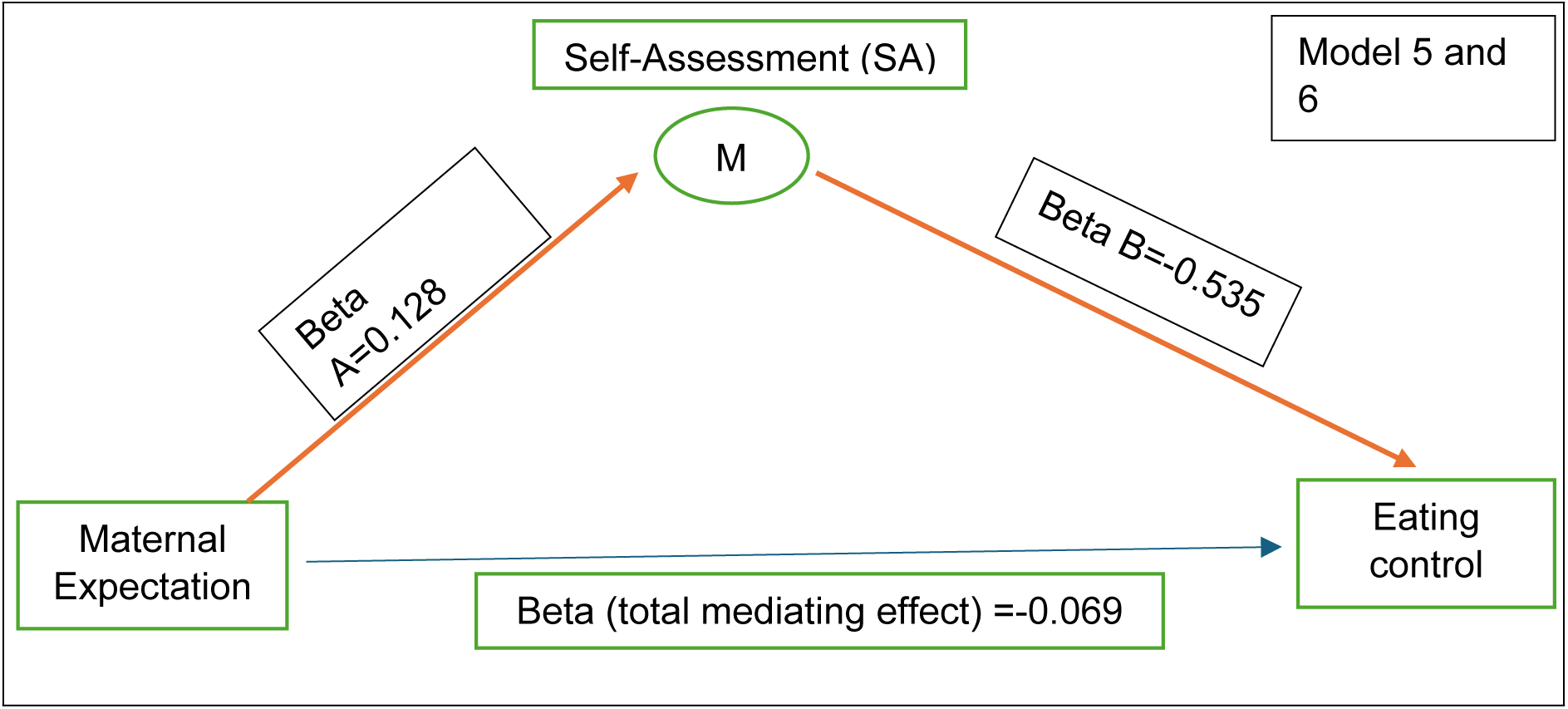
Mediating effect of Self-Assessment (SA) dimension on eating control in terms of maternal expectations (Red bold arrows illustrate the significant relationships)

Models 7 and 8 showed that self-assessment significantly and positively mediated the association between maternal control and dieting (Total mediation). There was no significant direct effect of maternal control on the dieting (Table 3; Figure 4).

**Figure 4.**
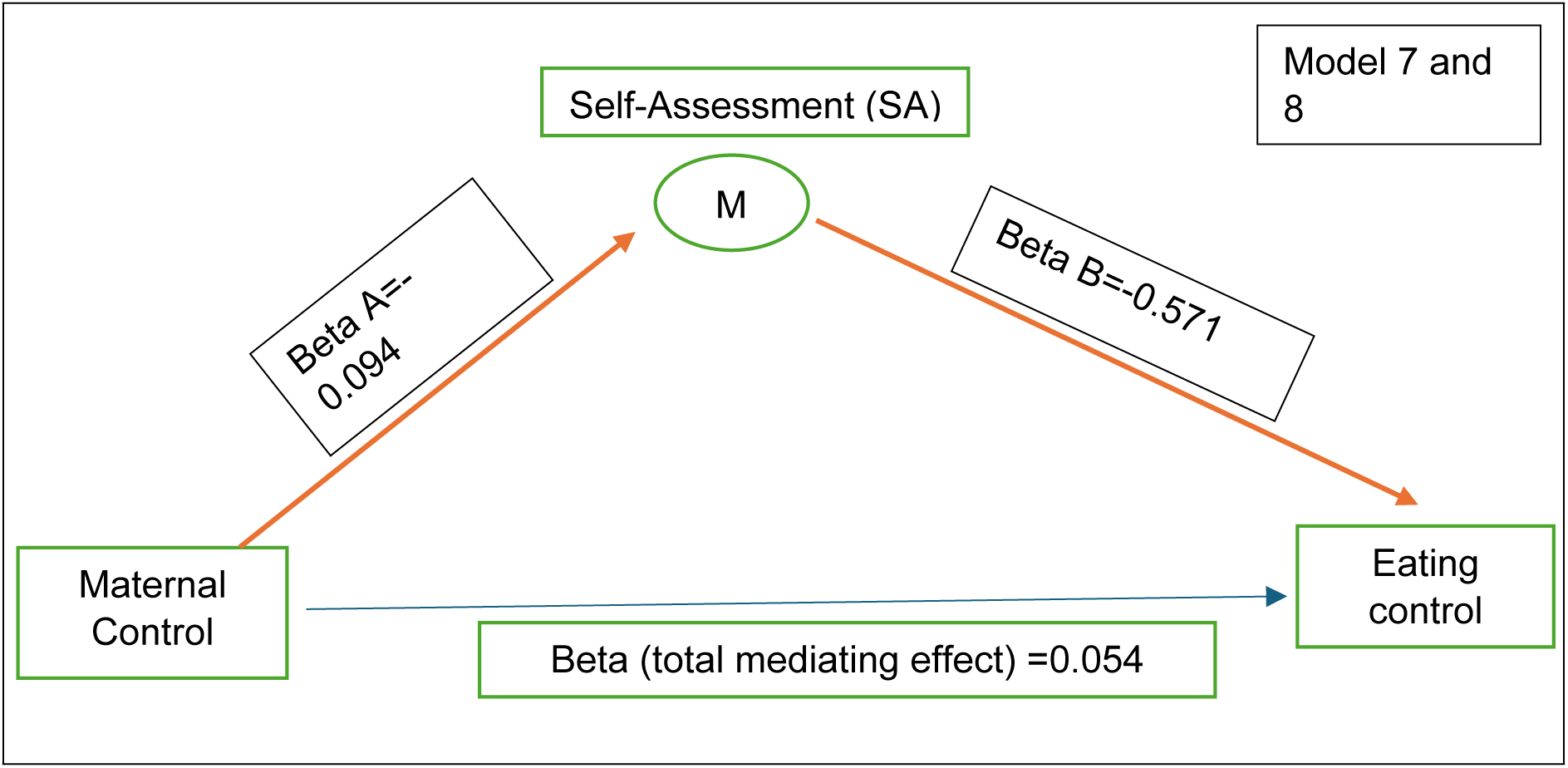
Mediating effect of Self-Assessment (SA) dimension on eating control in terms of maternal control (Red bold arrows illustrate the significant relationships)

Models 9 and 10 showed that self-assessment significantly and negatively mediated the association between maternal permissive and dieting (Total mediation). There was no significant direct effect of maternal permissive on the dieting (Table 3; Figure 5).

**Figure 5.**
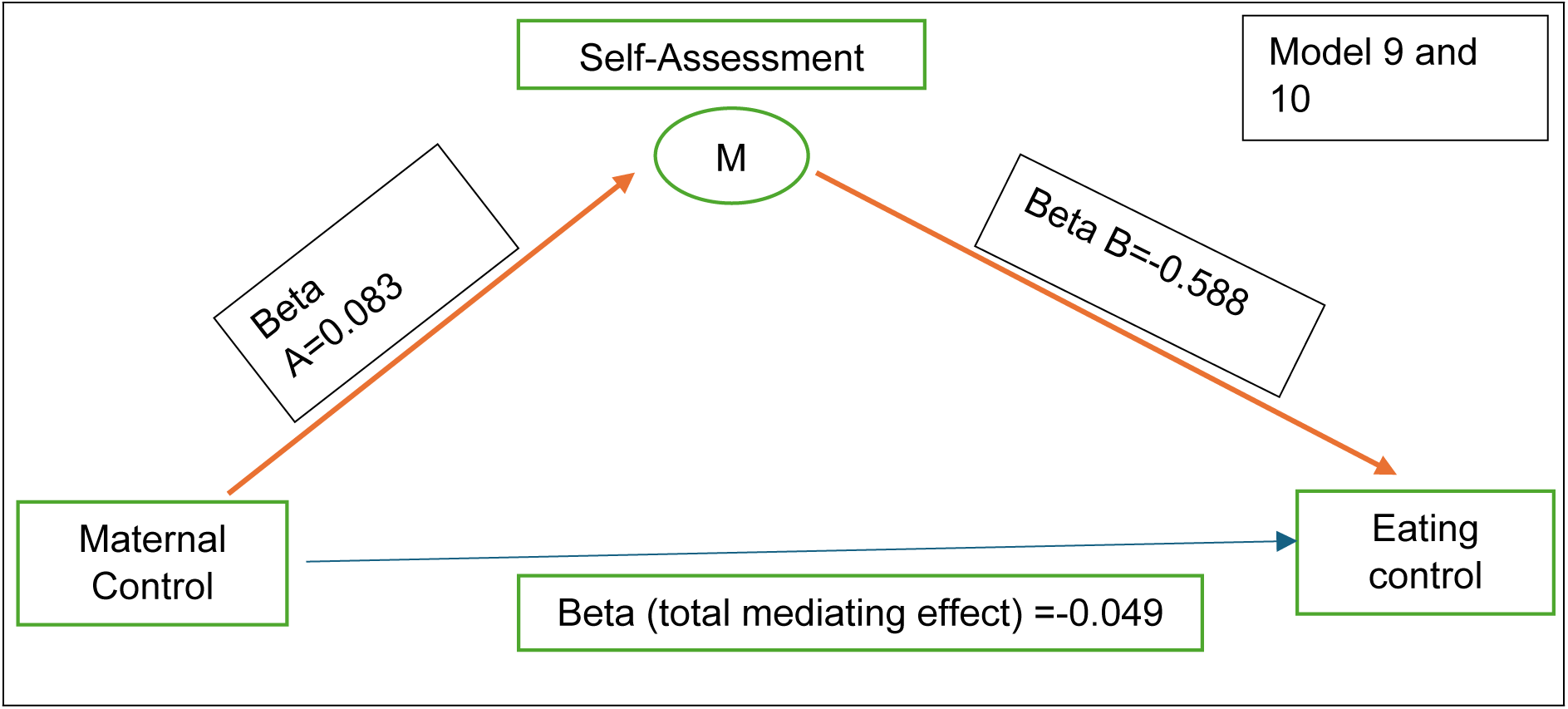
Linear regression of Self-Assessment (SA) dimension as dependent variable and maternal control as the independent variable (Red bold arrows illustrate the significant relationships)

#### Mediating effect of Self-Assessment on Bulimia Subscale

Models 11 and 12 showed that self-assessment significantly and negatively mediated the association between maternal expectations and bulimia (Total mediation). There was no significant direct effect of maternal expectations on the bulimia (Table 3; Figure 6).

**Figure 6.**
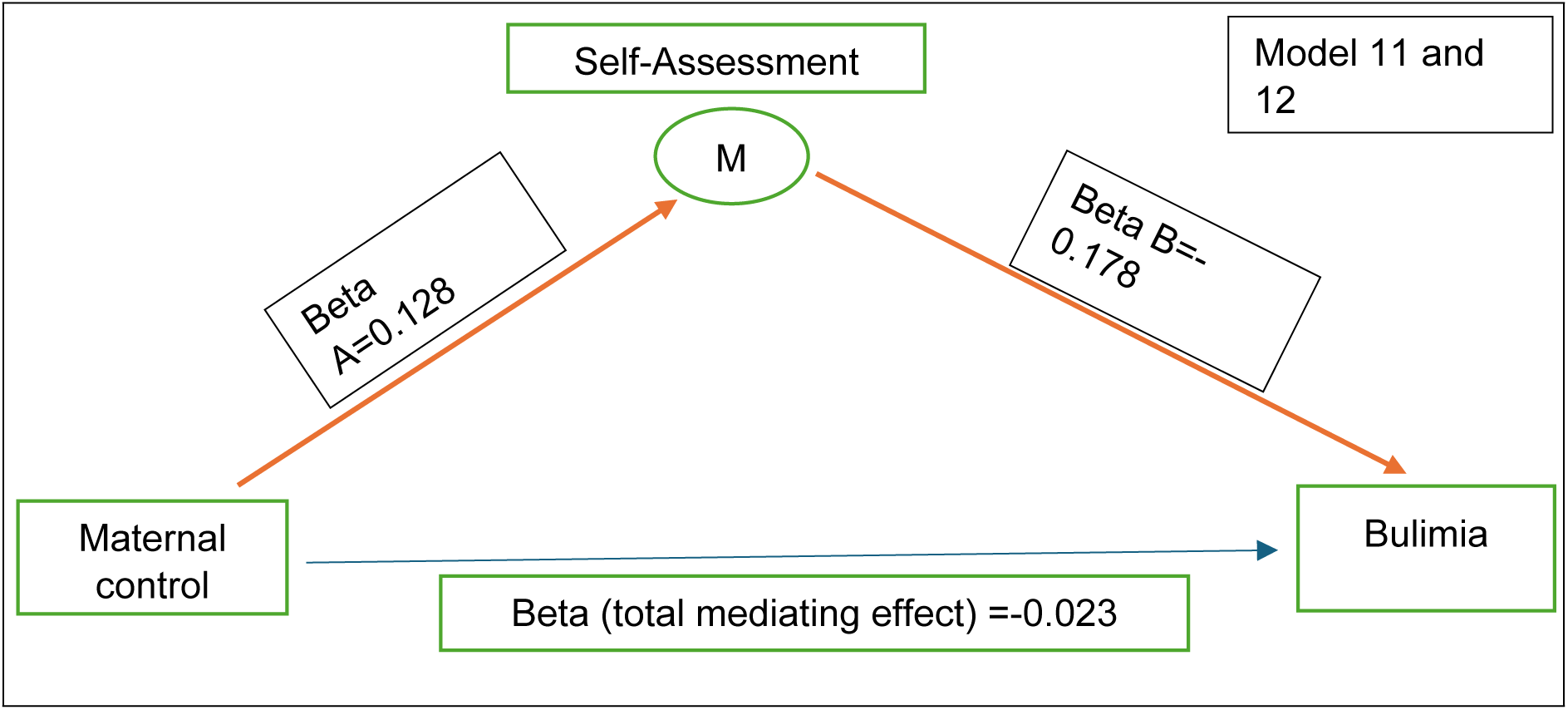
Mediating effect of Self-Assessment (SA) dimension on Eating dimension on bulimia in terms of maternal control (Red bold arrows illustrate the significant relationships)

Models 13 and 14 showed that self-assessment significantly and positively mediated the association between maternal control and bulimia (Total mediation). There was no significant direct effect of maternal control on bulimia. (Table 3; Figure 7).

**Figure 7.**
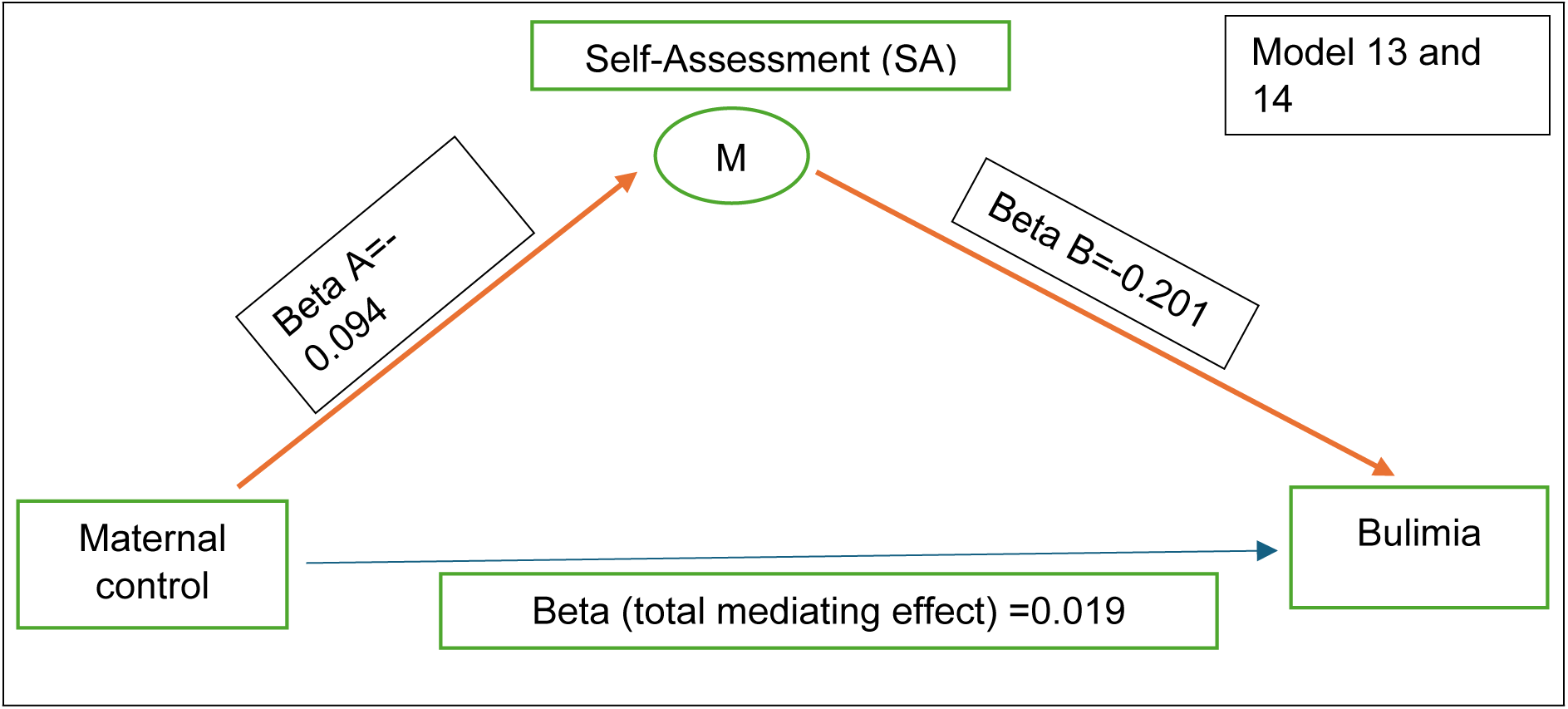
Mediating effect of Self-Assessment (SA) dimension on bulimia in terms of maternal control (Red bold arrows illustrate the significant relationships).

## Discussion

This study aimed to investigate the mediating role of self-assessment, one of the two dimensions of self-esteem, in the relationship between maternal parenting styles and eating disorders in a sample of Lebanese university students. We initially hypothesized that a person raised in more warmth and support from one’s mother, would exhibit fewer eating disorders because this relation would be facilitated by higher self-evaluation and positive body image. Our findings indicate a complex interplay between mothers parenting approaches and ED, whereby specific characteristics of mothers’ conduct may affect young adults’ eating behaviors, totally mediated by the self-assessment aspect of self-esteem.

In terms of mediational models, our study showed that permissive and highly expecting maternal style may be supporting to higher self-assessment (β=-0.049, p= 0.306, 95% CI between - 0.003 to 0.166). This finding is similar to previous research showing that permissive maternal styles lead to improved self-assessment (Zarychta et al.,2019). Authoritative maternal style was also found to lead to higher self-assessment causing a lower possibility of having an eating disorder, eating control, and dieting behaviors (β=-0.027, p<0.001, 95% CI between -0.800 to 0.039). This is in line with the literature highlighting that authoritative maternal style is correlated with lower disordered eating behaviors, better eating lifestyle, and improved self-assessment (Zohar et al.,2021). In addition, elevated maternal expectations increase self-assessment leading to decreased instances of disordered eating habits (β=-0.069, p=0.166, 95% CI between -0.246 and 0.042). This relationship, in line with previous research, may be explained by the possibility that children with authoritative parents may have better self-regulation abilities and self-efficacy, guarding them against poor eating habits (Rhee et al.,2013). Specifically, emotional support and positive reinforcement adopted by authoritative parents might improve self-assessment and reduce the vulnerability of children to disordered eating as a means of gaining control (Rhee et al.,2013). This finding is consistent with another study indicating that favorable mental health outcomes, such as internalizing self-regulation and healthy eating habits, are linked to authoritative parenting styles, which are typified by high warmth and appropriate control (Zohar et al., 2021). In a similar vein, the hypothesis of authoritative parenting’s preventive role against ED is supported by the present results. In addition, the current finding aligns well with the social learning theory (Bandura, 1986), which holds that healthy eating habits and a lower risk of disordered eating can be shaped by positive behaviors such as self-regulation that maternal figures model.

However, high maternal control was found to cause lower self-assessment leading to higher eating disorders (β=0.054, p=0.224, 95% CI between -0.064 and 0.271). Previous research showed that higher eating disorders are associated with high maternal control which is similar to our study (Scotto et al.,2024). The eating control and dieting were correlated with lower self-assessment indicating that students who have controlled strict eating behaviors can lead to ED, particularly if the maternal style is unsupportive, less permissive, and less responsive (Zarychta et al.,2019). A higher bulimia level was correlated with lower self-assessment and higher maternal expectations (β=-0.023, p=0.146, 95%CI between -0.108 and 0.016) and control (β=0.019, p=0.467, 95%CI between -0.045 and 0.099). our comparison to the literature reveals that bulimia may increase with higher maternal expectations and control because they may be unsupportive, lead to low self-assessment, and cause disordered eating behaviors (Cascino et al,2023).

This result is in line with other studies showing that ED is more likely to develop in homes with high levels of parental control and authoritarian parenting (Hampshire et al., 2022; Usmani et al., 2022). Similarly, previous work indicates that emotional overeating and disordered eating habits are more likely in cases of excessive maternal control, particularly when combined with insufficient emotional care (Monteleone et al., 2020). One possible explanation might be that children who experience excessive parental control may internalize emotions of worthlessness or failure, encouraging disordered eating as a maladaptive coping strategy (Farrow et al.,2015; Hampshire et al.,2022). In turn, the findings of previous research have been nuanced by the present study, which emphasizes the role that self-assessment plays as a crucial mediator in this connection (Messeri et al.,2023; Mora, 2017). Our study also suggests that excessive control weakens self-assessment (β=0.019, p=0.003, 95%CI between -0.156 and -0.032) which may help to explain why it leads to disordered eating. This emphasizes the value of parents having reasonable expectations for their children. While setting high expectations may drive children to excel and reveal positive outcomes, these expectations may impose significant pressure on the child and potentially affect their mental health, especially if excessive control is without warmth (Messeri et al.,2023; Zohar et al., 2021).

### Clinical implications

Positive clinical implications stem from the present study’s findings, especially regarding the prevention and treatment of ED in university students in Lebanon. Improved self-esteem therapies might serve as an effective preventative strategy due to the critical role that self-esteem plays as a mediator between maternal parenting styles and disordered eating habits. For example, while dealing with families, psychologists should stress the need for balanced parenting that promotes self-control and constructive self-evaluation. Parenting courses and family-based treatment may also be devised to teach parents—especially mothers—the advantages of authoritative parenting and how to avoid the dangers of overcontrol, which lowers self-esteem and raises the likelihood of ED (Harrer et al., 2020). In parallel, programs that enhance emotional control and self-evaluation may also help lower the likelihood of ED. Furthermore, the findings are not only restricted to the practice of private clinicians but also extend to those working closely with universities. As such, university physicians might conduct preventative initiatives aimed at helping students develop emotional fortitude and self-worth. By addressing the effects of family dynamics, media influences, and self-perception, workshops, and peer support groups may lessen the risk of ED in students by assisting them in navigating the demands of body image and academic stress (Forbush et al., 2023).

### Strengths and limitations

While this study provides novel data and makes important strides by examining the mediation effect of self-esteem on the association between parenting style and eating disorders, it is not without limitations. The cross-sectional design precludes the ability to establish causality, and our methodology does not fully eliminate the potential for selection or information bias, which could lead to an underestimation of the findings. Despite these challenges, the study holds strengths such as a robust sample size, rigorous standardized analysis, and a theoretical framework grounded in “the social cognitive theory” applied within complex models. Future research, either prospective or interventional, is needed to confirm and extend our findings.

### Future research

Future research is needed to further analyze the previously mentioned mediating variables with a specific focus on paternal parenting styles. Combined with the present knowledge, future research on paternal parenting styles would provide a more nuanced understanding of the existing body of literature. Additionally, future research could consider the examination of other social or cultural mediating variables. For instance, the examination of media exposure as a mediating variable may add layer to the present understanding between parenting styles and ED.

## Conclusion

This study mostly provides healthcare professionals and clinical educators with valuable insights into the complex association between parenting and ED and highlights the importance of developing skillsets to boost self-esteem for those at risk of developing ED.

### Public Significance Statement

This study shows the complexity of the relation between mothers parenting styles, self-esteem of offsprings and eating disorders in Lebanese university students. Findings highlight the need to help mothers understand the impact of their connection to their kids on eating patterns and also highlight the need for interventions targeting self-esteem, particularly self-assessment, to prevent and subsequently potentially treat eating disorders, informing clinical practice and educational programs for healthcare professionals and families.

## Data Availability

Data are available from the LAU Repository for researchers who meet the criteria for access to confidential data.

## Ethical statement

The study received ethical approval by the Lebanese American University- Institutional Review Board (LAU-IRB) (LAU IRB: LAU.SAS.TS1.10/Nov/2023). Before completing the questionnaire, all participants provided their written consent by clicking on the respective button on the first page of the Google Form.

## Author contributions

TS and MEM designed research; TS and MEM reviewed the manuscript; TH and JM collected data; SBM analyzed data; TH, SBM, and RR wrote the paper. All authors read and approved the final manuscript.

## Data Availability

Data will be made available upon reasonable request to authors

## Permission to reproduce material from other sources

N/A

